# De-medicalising gluten-free products through a subsidy card scheme: a qualitative study of service users

**DOI:** 10.1101/2024.12.09.24318385

**Authors:** Abubakar Sha’aban, Francesca Mazzaschi, Beti-Jane Ingram, Natalie Joseph-Williams

**Affiliations:** Health and Care Research Wales Evidence Centre, Cardiff University, United Kingdom; Health and Care Research Wales Evidence Centre Public Partnership Group, Cardiff University, United Kingdom

**Author notes:** **Funding Statement** The authors and their Institution were funded for this work by the Health and Care Research Wales Evidence Centre, itself funded by Health and Care Research Wales on behalf of Welsh Government.

## Abstract

**Introduction:** Coeliac disease requires a strict gluten-free diet to prevent complications. In the late 1960s, Gluten free foods (GFF) became available on prescription to support adherence when these foods were scarce in shops. Today, gluten-free foods are more accessible in large supermarkets, making it easier for patients to obtain them without prescriptions. In 2019, Hywel Dda University Health Board introduced the GFF subsidy card as an alternative to prescriptions, a method still used by other health boards in Wales.

**Aims:** This study aimed to explore individuals’ perspectives on a new subsidy card scheme for GFF, identify key barriers and facilitators, and provide recommendations to enhance the scheme’s implementation and effectiveness.

**Method:** A qualitative approach was employed, involving in-depth interviews with coeliac patients and parents of coeliac children. Participants included current users of the subsidy card, those relying on prescriptions, and individuals who opted out of the card scheme.

**Results:** Participants using the subsidy card valued its flexibility, variety of products, and convenience over the prescription system. Challenges included managing card balances, limited retailer acceptance, and geographical disparities. Non-users appreciated the potential for increased choice and mostly showed interest in switching but some raised concerns about inflation, misuse, and the additional burden on taxpayers.

**Conclusion:** Most participants view the subsidy card as an acceptable alternative to GFF prescriptions. To improve the subsidy card uptake, recommendations include enhancing digital tools for balance management, card value reviews, expanding retailer partnerships, promoting safer food practices, targeted communication, and ensuring prescription continuity for those who prefer it. These measures can optimize the scheme’s impact and better support individuals with coeliac disease.

## Background

Coeliac disease is an autoimmune condition requiring a lifelong gluten-free diet. In Wales, gluten-free foods are prescribed, but pre-paid subsidy cards are being explored as a cost-effective alternative. A 2018 pilot by Hywel Dda University Health Board involved 123 participants across nine GP practices, with 86% preferring the card over prescriptions due to greater choice and convenience. However, some opted out, citing limited financial benefits and product availability. The subsidy cards scheme was fully implemented by the Hywel Dda board in 2019, but further research is needed to assess its impact on patients’ quality of life, its economic implications, and the barriers faced by those who continue to use prescriptions.

### The aim of this study

The aim of this study is to explore service users’ views on the use of pre-paid subsidy cards in place of prescriptions, including both those who were invited to take part in the HDUHB pilot and those who have and have not been offered to participate in the scheme to date.

Specific objectives were to explore:

- **experiences** of current services received (prescription or subsidy card)
- **impact** of current methods of accessing GFF on daily life
- perceived **benefits and disadvantages** of switching to a pre-paid subsidy card
- **barriers and facilitators** to participation in a pre-paid subsidy card scheme
- **main priorities** regarding access to gluten-free products – what matters most to individuals?
- **recommendations** for future roll-out
- **reasons for declining** to participate (for those who declined to participate in the HDUHB pilot scheme)

## Methods

This qualitative study used semi-structured Zoom interviews (April–July 2024) to explore views of individuals eligible for the subsidy card scheme in HDUHB and those potentially eligible in a future Wales-wide roll-out. Researchers followed an interview schedule to ensure key topics were covered, with all interviews recorded.

### Key Findings

This study included 29 participants from all seven health boards in Wales. Of these, 23 were personally eligible for gluten-free food (GFF) provision, while six managed GFF for others.

Participants from health boards without the subsidy card scheme and two from Hywel Dda University Health Board (HDUHB) who declined the card, highlighted benefits of prescriptions, including financial savings, guaranteed access to essentials, and convenience. However, issues like complicated ordering, product unavailability, and food waste were drawbacks.

Among HDUHB participants in the subsidy card scheme, most praised its flexibility, increased product variety, and ease of use, citing reduced financial strain and enhanced social inclusion. Some challenges included card restrictions, difficulty checking balances, and limited rural access to GFF products.

Participants from other health boards expressed interest in the card’s benefits, such as improved dietary options, but raised concerns about its value in the current economic climate, potential misuse, and rural access barriers. Most (70%) were interested in switching to the subsidy card, while a minority expressed ambivalence or reluctance.

### Key Priorities

Six key priorities for improving access to GFF and better supporting people with coeliac disease were identified:

1. **Ease of Access**
2. **Variety & freedom of choice**
3. **Tasty and healthy diet**
4. **Consistency**
5. **Cost**
6. **Better awareness & education of coeliac disease**

## 1. Background

Coeliac disease is an autoimmune condition that causes chronic inflammation in the small intestine. Dietary proteins known as gluten, which are present in wheat, barley and rye, activate this inflammation resulting in the malabsorption of nutrients. The management of coeliac disease is aimed at eliminating symptoms (such as diarrhoea, bloating and abdominal pain) and reducing the risk of complications, including those resulting from malabsorption and following a gluten-free diet is the only effective management for the condition. Unlike other foodstuffs, gluten containing products are not necessary for a healthy diet and patients with gluten sensitivity can safely exclude it from their diet and still eat healthily without needing special foods, but these are available for those who require them.

In Wales, gluten free foods are available to purchase and those diagnosed with coeliac disease (and those with a diagnosis of Dermatitis Herpetiformis) can receive gluten free food on prescription in accordance with the national prescribing guidelines (AWMSG, 2018). However, there is increasing interest in the potential benefits of pre-paid subsidy cards for people with coeliac disease as an alternative to prescription supply.

In 2018, the Hywel Dda University Health Board published a service evaluation report on a opt-in pilot scheme, where pre-paid subsidy cards were used to compensate individuals for the higher costs of purchasing gluten-free products, in place of prescription (Jones, 2019). The cost of providing gluten-free products via prescription is generally at a greater cost to the NHS compared to purchase through retail outlets. Therefore, it is important to understand whether supporting patients to purchase gluten-free products would be a favourable alternative to prescriptions in terms of quality of care, experiences of accessing gluten free food (GFF), and economic benefits for patients; as well as reduced cost to the NHS (DHSC, 2018).

Nine GP practices were involved in this initial pilot scheme, and 123 of the 195 (63%) service users approached agreed to participate. Participants received a pre-paid subsidy card, which included a monetary amount determined to cover the extra costs incurred when purchasing specialised gluten-free products. The results of the service evaluation questionnaire indicated that the use of a subsidy card was preferred over prescriptions, and 86% of participants opted to continue using the card at the end of the pilot.

Benefits of the card reported by the participants included improved choice, avoiding the need for large stocks of produce, and an increased sense of control and normalisation of their dietary needs. However, whilst the card was generally preferred, 17 (14%) participants decided to not continue using the card; they felt it did not benefit them financially and some products were still only available through prescription, meaning they could not access the products they wanted with the card.

Following the positive feedback from the pilot, Hywel Dda University Health Board expanded the scheme across the Health Board. It is now hoped that this will be extended to Health Boards across Wales. Whilst annual evaluations using surveys to understand the acceptability of this scheme have been conducted by the Hywel Dda University Health Board (HDUHB), there are limited data available on the impact the card scheme has on service users’ day to day lives, their quality of life and the perceived economic implications. Further, there is a lack of understanding of the motivations of those who wish to remain using prescriptions, and we also know little about the views of those who have not yet been offered to participate in the scheme. Establishing a better understanding of this could help identify barriers to participation and facilitators to uptake of the scheme and help guide improvement and future roll-out of the subsidy card scheme.

## 2. Aims and Objectives

The aim of this study is to explore service users’ views on the use of pre-paid subsidy cards in place of prescriptions, including both those who were invited to take part in the HDUHB pilot and those who have and have not been offered to participate in the scheme to date.

**Key objectives** included exploring:

- **experiences** of current services received (prescription or subsidy card)
- **impact** of current methods of accessing GFF on daily life
- perceived **benefits and disadvantages** of switching to a pre-paid subsidy card
- **barriers and facilitators** to participation in a pre-paid subsidy card scheme
- **main priorities** regarding access to gluten-free products – what matters most to individuals?
- **recommendations** for future roll-out
- **reasons for declining** to participate (for those who declined to participate in the HDUHB pilot scheme)

## 3. Methods

This was a qualitative study using semi-structured interviews to explore the views of those who are currently eligible for the subsidy card scheme in the HDUHB and those who would be eligible to part of the scheme in a potential future roll-out across other health boards in Wales.

Ethical approval for this study was granted by Cardiff University School of Medicine Research Ethics Committee (SMREC 24.21). Recruitment processes and documents were developed with our Public Partner (B-JI) to ensure they were feasible, understandable, and relevant.

### 3.1 Participant recruitment

Participants included all Welsh residents who are eligible to receive GFF through NHS prescriptions; for participants based in HDUHB, it included those who were eligible for a pre-paid subsidy card for the purchase of GFF. Eligibility criteria for the study are presented in ***Table 1***. Participants were recruited through social media ‘X’ (formerly known as Twitter) accounts of Health and Care Research Wales, Health and Care Research Wales Evidence Centre, and The Wales Centre for Primary and Emergency Care Research (PRIME Centre Wales). We also recruited through relevant Facebook accounts for people with coeliac disease. The opportunity was also shared via the Coeliac UK (a charity for people who need to live without gluten) mailing list to members who live in Wales.

**Table 1.**
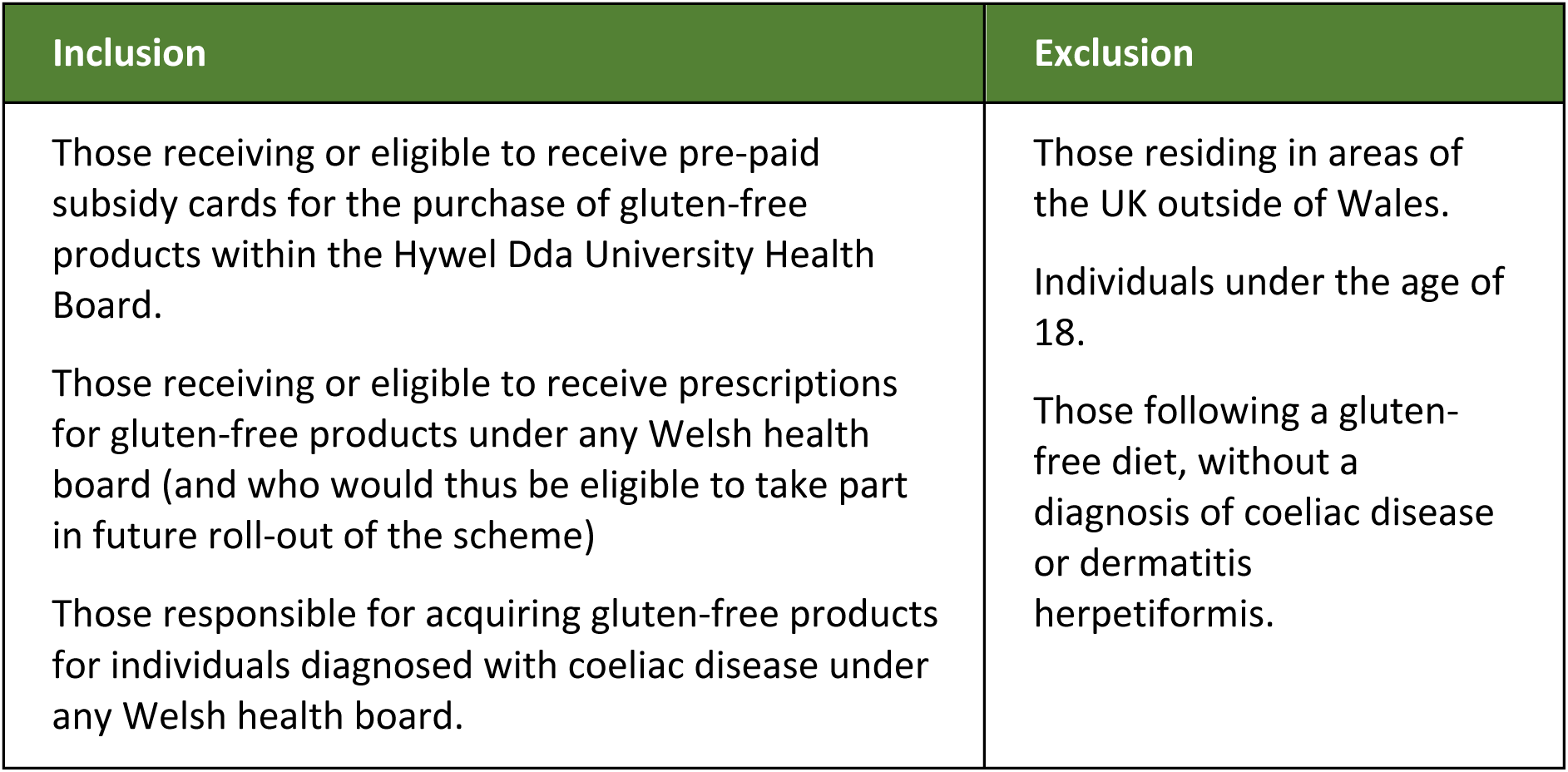
Eligibility Criteria.

Advertisements included a poster summarising the study (Appendix A) and a link to the study webpage, which included a detailed participant information sheet (Appendix B). Interested participants were asked to complete an online screening and demographic survey (Appendix C) to register their interest in taking part. Demographic variables collected included: gender, age group, health board area, eligibility for NHS gluten-free food provision in Wales, gluten-free food access method, disability status, highest educational level, employment status, personal income range (previous year), and ethnicity. We also asked participants three questions exploring components of digital literacy (Nelson, Pennings, Sommer, Popescu, & Barkin, 2022). We used data from the screening and demographic survey to purposively select a diverse and representative sample. If selected, participants were notified via email and asked to complete a consent form (Appendix D); a convenient date and time were agreed upon for the interview with the researcher (AS/FM).

### 3.2 Data collection

Semi-structured interviews were conducted online via Zoom between April 2024 and July 2024. Interviews were conducted by AS or FM and audio/video recorded. Researchers (AS/FM) used an interview schedule (Appendix E) to guide the discussion and ensure key topics were explored. This was developed with stakeholders, including our Public Partner (B-JI), to ensure the questions were accurate, accessible, and covered areas that mattered to Coeliac patients. Although a single interview schedule was used, there were sections with questions tailored to specific participant circumstances, namely:

- Section 3a: For participants who were offered the card scheme in Hywel Dda and accepted it
- Section 3b: For participants who were offered the card scheme in Hywel Dda and declined it
- Section 3c: For participants from other local health boards where the scheme is not yet available

A summary of key topics covered in the interview guide is included in Box 1.

#### Box 1

**Summary of key topics explored in the Interview guides**

- Experiences of current services received
- Impact of current methods of accessing GFF on daily life
- Perceived benefits or disadvantages of using prescriptions to access gluten-free products
- Perceived benefits or disadvantages of using/switching to a pre-paid subsidy card system
- Main priorities regarding access to gluten-free products – what matters most to individuals?
- Suggestions for improving access to gluten-free products
- Recommendations for future roll-out of the subsidy card scheme

### 3.3 Data Analysis

Interviews were transcribed verbatim for analysis using a Cardiff University authorised third-party transcription service. Transcripts were imported into NVivo 12 pro Qualitative Analysis Software (Lumivero, 2017) for analysis. Thematic analysis (Braun & Clarke, 2006) was applied to the data (Gale, Heath, Cameron, Rashid, & Redwood, 2013) by AS and FM; this is a systematic method used to identify patterns and themes within data. It involves a series of structured steps including familiarisation with the data, coding, developing a thematic framework, charting the data into the framework (including verbatim quotes), and interpreting the key themes.

The initial thematic coding framework (emerging themes from preliminary exploration of the data) were applied to a set of transcripts (n=6) by FM and cross-checked by AS. The emerging frameworks were discussed with NJW and refined (similar codes merged and additional codes added to the framework) before applying the frameworks to the remainder of the transcripts. The data is presented in the key findings section (see section 4.2).

## 4. Results

### 4.1 Participants

We recruited a total of 29 participants across all health boards. Overall, 79% (n=23) of participants identified as female; 21% identified as male (n=6). Participants reported a range of disability status, educational attainment, employment status, and personal income range (see ***Table 2***).

**Table 2.** Demographic characteristics of study participants (n=29)

| Research ID | GFF Access Method | Gender<br>M = Male<br>F = Female | Personal eligibility<br>for NHS GFF<br>Provision in Wales | Personal income last year (£) |
| --- | --- | --- | --- | --- |
| **Hywel Dda University Health Board (n=6)** |  |  |  |  |
| P01 | Pre-paid subsidy card | F | Self-Eligible for GFF | £50, 000 to £74, 999 |
| P06 | Pre-paid subsidy card | M | Self-Eligible for GFF | £50, 000 to £74, 999 |
| P17 | Pre-paid subsidy card | F | Self-Eligible for GFF | £25, 000 to £49, 999 |
| P25 | Pre-paid subsidy card | F | Self-Eligible for GFF | £1 to £9, 999 |
| P09 | Prescriptions | F | Self-Eligible for GFF | £10, 000 to £24, 999 |
| P15 | Prescriptions | M | Self-Eligible for GFF | £25, 000 to £49, 999 |
| **Other Health Boards (n=23)** |  |  |  |  |
| P03 | Prescriptions | F | Self-Eligible for GFF | £25, 000 to £49, 999 |
| P05 | Prescriptions | F | Self-Eligible for GFF | £25, 000 to £49, 999 |
| P07 | Prescriptions | F | Self-Eligible for GFF | £25, 000 to £49, 999 |
| P08 | Prescriptions | F | Self-Eligible for GFF | £10, 000 to £24, 999 |
| P11 | Prescriptions | F | Self-Eligible for GFF | £10, 000 to £24, 999 |
| P12 | Prescriptions | F | Self-Eligible for GFF | £50, 000 to £74, 999 |
| P13 | Prescriptions | M | Self-Eligible for GFF | £1 to £9, 999 |
| P14 | sole purchaser | F | Self-Eligible for GFF | £25, 000 to £49, 999 |
| P18 | Prescriptions | F | Self-Eligible for GFF | £10, 000 to £24, 999 |
| P21 | Prescriptions | F | Self-Eligible for GFF | £25, 000 to £49, 999 |
| P22 | Prescriptions | M | Self-Eligible for GFF | £25, 000 to £49, 999 |
| P23 | sole purchaser | F | Self-Eligible for GFF | £25, 000 to £49, 999 |
| P24 | Prescriptions | F | Self-Eligible for GFF | £25, 000 to £49, 999 |
| P26 | sole purchaser | F | Self-Eligible for GFF | £10, 000 to £24, 999 |
| P27 | sole purchaser | F | Self-Eligible for GFF | £10, 000 to £24, 999 |
| P28 | Prescriptions | F | Self-Eligible for GFF | £10, 000 to £24, 999 |
| P29 | Prescriptions | F | Responsible for an Eligible Child | £50, 000 to £74, 999 |
| P02 | Prescriptions | F | Responsible for an Eligible Child | £25, 000 to £49, 999 |
| P04 | Prescriptions | F | Responsible for an Eligible Child | £10, 000 to £24, 999 |
| P10 | Prescriptions | M | Responsible for an Eligible Spouse | £10, 000 to £24, 999 |
| P16 | Prescriptions | M | Responsible for an Eligible Child | £50, 000 to £74, 999 |
| P19 | Prescriptions | F | Responsible for an Eligible Child | £25, 000 to £49, 999 |
| P20 | Prescriptions | F | Responsible for an Eligible Child | Prefer not to say |

#### 4.1.1 Local Health Board Area

All seven health boards in Wales were represented by participants in the interviews (See ***Figure 1***); 21% (n=6) of participants lived in the Hywel Dda University Health Board area and 23 participants from other Health Boards interviews (See ***Figure 1***).

**Figure 1 -.**
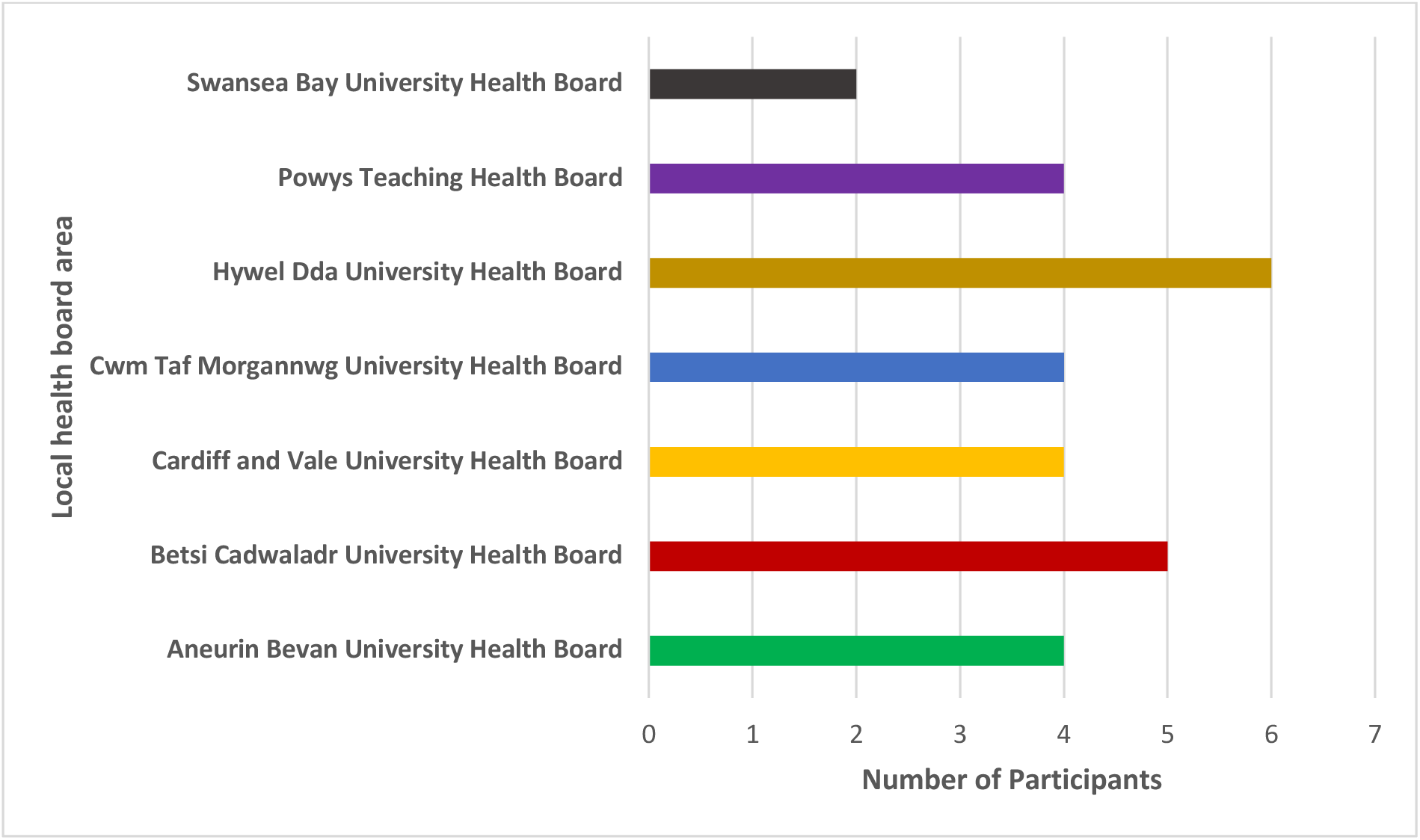
Local Health Board area of participants.

#### 4.1.2 Personal eligibility for NHS gluten-free food provision in Wales

Of the 29 study participants, 23 (79%) are personally eligible for gluten-free food (GFF) provision in Wales, while 6 (21%) are responsible for individuals eligible to receive gluten-free food through the NHS.

#### 4.1.3 Gluten-free food access method

Among the six participants from HDUHB (who were part of the subsidy card pilot scheme), four access their GFF through a pre-paid subsidy card. Two were offered the card but declined, opting to continue accessing their GFF through prescriptions. Of the 23 participants from other health boards who are eligible for prescriptions, four choose not to use them. See **Figure 2** for details.

**Figure 2 -.**
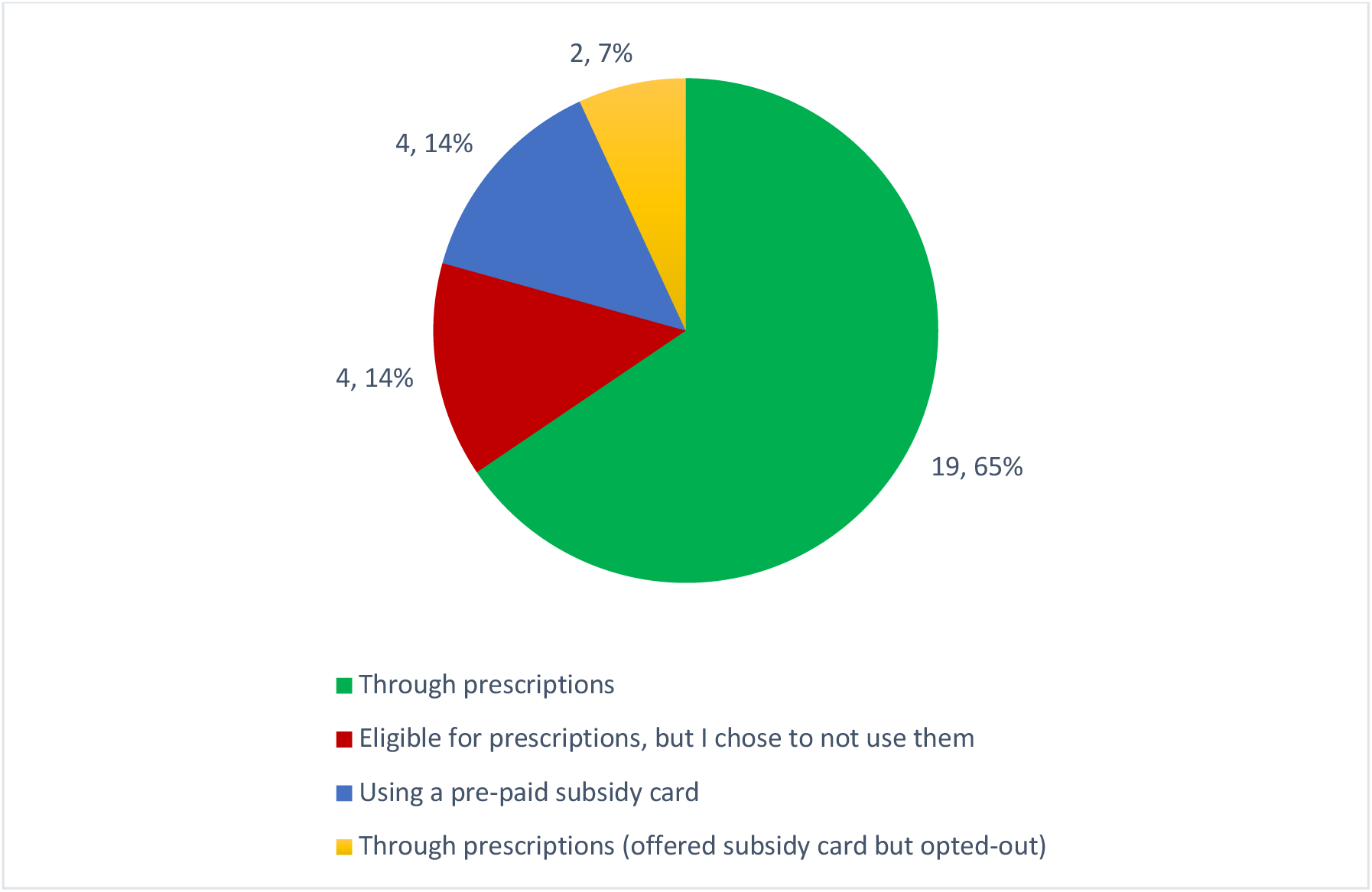
Number of participants accessing GFF through different methods.

#### 4.1.4 Age

Participants age groups ranged from 18–24 to 75–84, with the most frequent age categories being 35–44 and 55–64. (see ***Figure* 3**).

**Figure 3 -.**
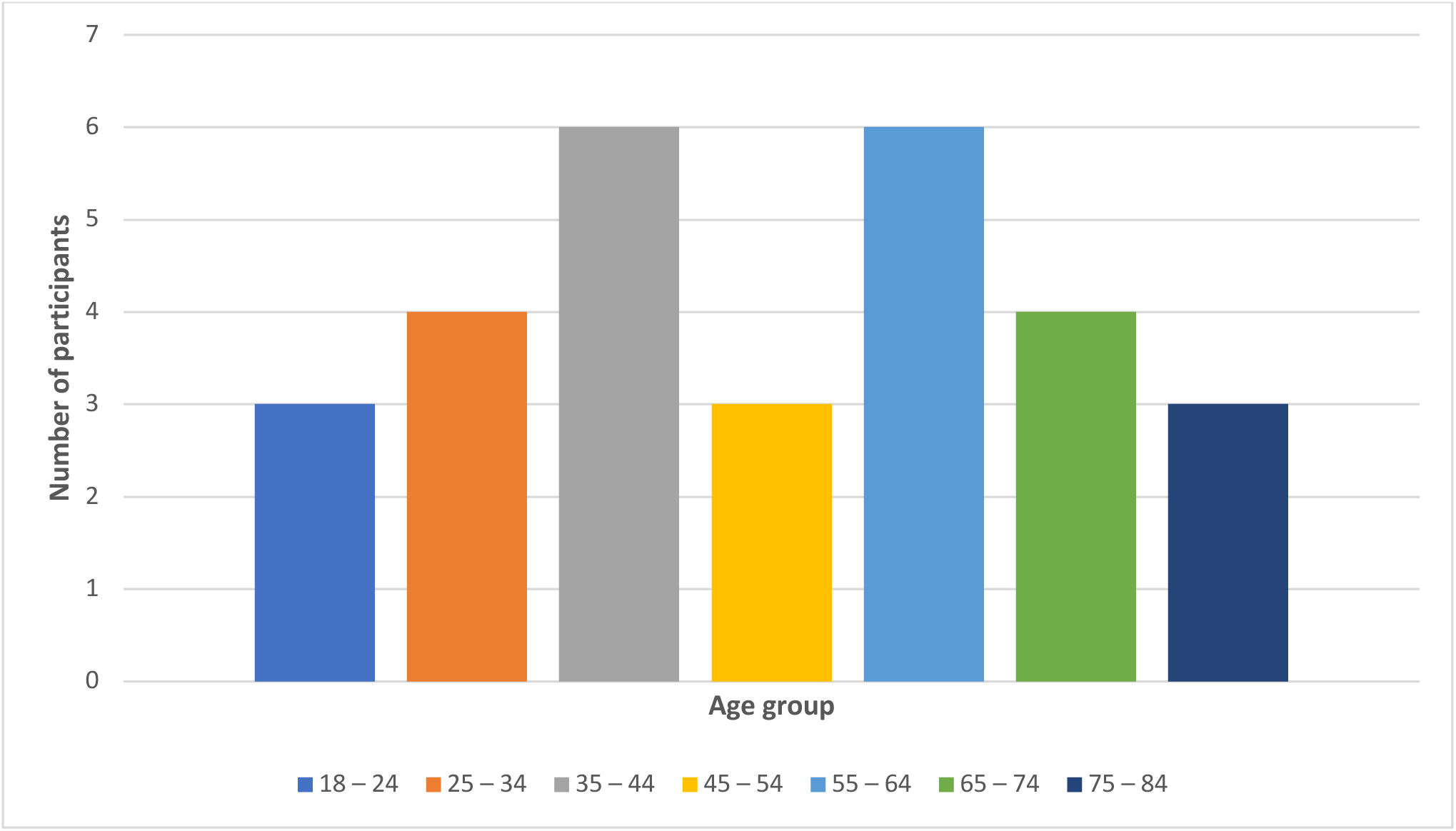
Age categories of participants.

#### 4.1.5 Digital Literacy

Participants reported a range of self-reported ‘digital literacy’, covering the use of applications, setting up video calls, and solving basic technical issues (see **Figure *4***).

**Figure 4 -.**
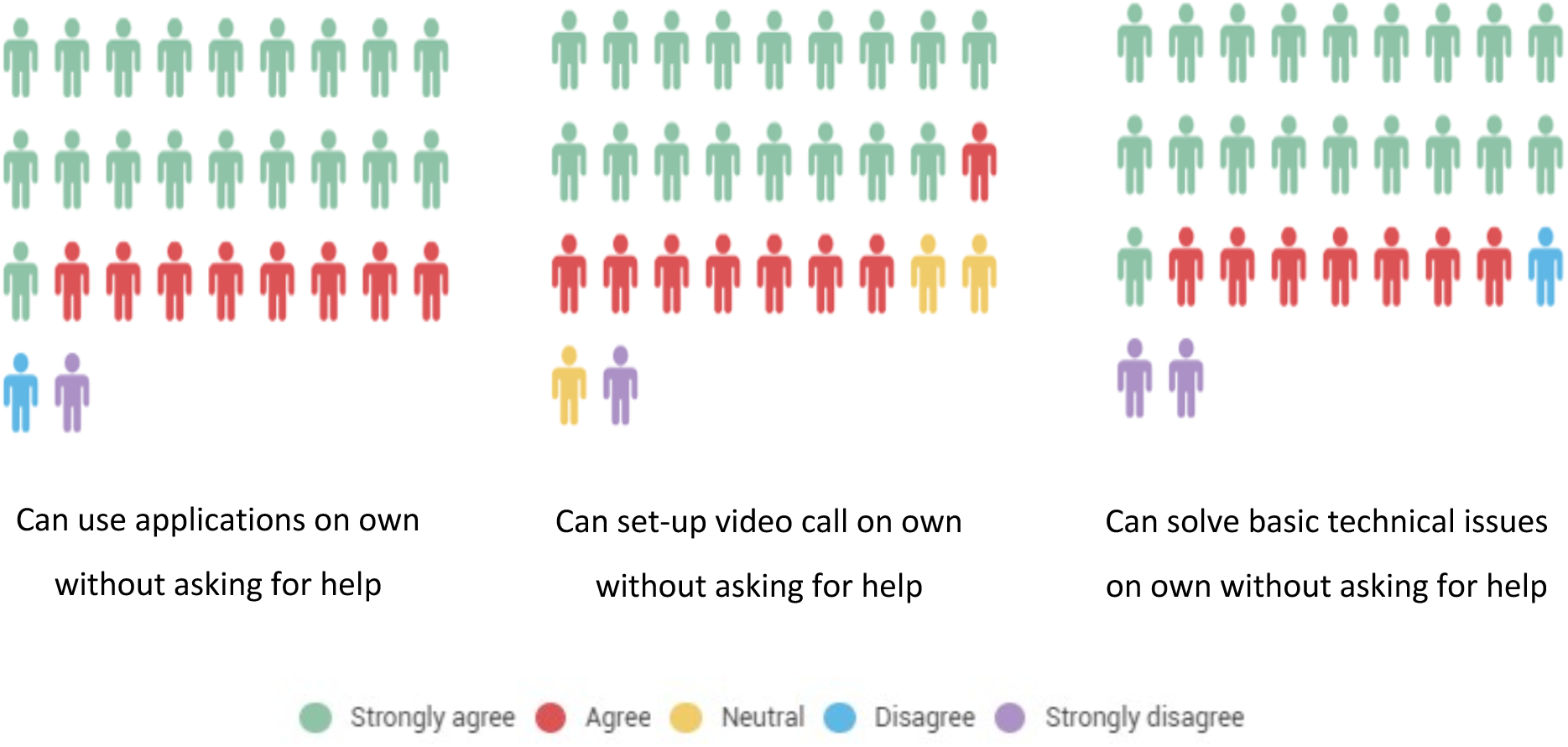
Self-reported level of digital literacy.

##### Use of applications

Overall, 27 participants (93%) agreed or strongly agreed that they could use applications/programs (like Zoom) on their mobile phone, computer, or another electronic device (for example a tablet) on their own without asking for help from others. Two participants disagreed or strongly disagreed.

##### Setting up video calls

Overall, 26 participants (90%) agreed or strongly agreed that they could set up video calls on their mobile phone, computer, or another electronic device on their own without asking for help from others. Three participants disagreed or strongly disagreed.

##### Solving basic technical issues

Overall, 25 participants (86%) agreed or strongly agreed that they could solve or figure out how to solve basic technical issues on their own without asking for help from others. One participant strongly disagreed and three were neutral.

### 4.2 Key Findings

Overall, six key themes emerged from the interviews. These themes are summarised in ***Table 3***. Detailed themes and sub-themes, along with exemplar quotes, are provided in Additional Table A (separate file) and are discussed in detail below. We analysed all the data collectively. However, some themes were specific to current users of the subsidy card scheme, others were relevant to individuals not yet using the scheme, and some themes were applicable to both groups. We note this when describing the theme.

**Table 3.** Summary of key themes and sub-themes.

| Theme | Report Section | Page Number |
| --- | --- | --- |
| Views on prescriptions | 4.2.1 | 21 |
| Subsidy card – views of pilot scheme services users | 4.2.2 | 24 |
| Subsidy card – views of service users in other health boards | 4.2.3 | 28 |
| Experiences of coeliac disease health care services in Wales | 4.2.4 | 33 |
| General views and experiences | 4.2.5 | 34 |
| Main priorities of people with coeliac disease | 4.2.6 | 39 |

#### 4.2.1 Views on prescriptions

The twenty-three participants from the six health boards where the subsidy card scheme is not yet available, along with two participants from HDUHB who declined to use the subsidy card, shared their views on the existing prescription access to GFF.

##### 4.2.1.1 Perceived advantages

Several key benefits of using prescriptions to obtain GFF were reported by participants. One significant advantage is the **financial savings** associated with prescriptions, over no prescriptions. Participants appreciate the cost relief provided by receiving gluten-free products for free, which would otherwise be expensive:

> “I think the most beneficial thing about the prescriptions is the fact that I’m accessing these products for free, whereas they would be quite expensive otherwise, and I do think the products I receive are quite high quality as well. So I think that’s the key benefits.” (P11_Prescription User)

Another benefit is the **specialised options** that prescriptions can offer. For example, some types of specialist flour are not available in regular stores but can be obtained through prescriptions:

> “The flour, because you can’t buy Juvela flour [laughter] I think that’s my only thing, is that there’s some things on prescription that you can’t get in the shops.” (P12_Prescription User)

The **assurance of receiving essentials** regardless of financial situation is also a key advantage. Participants’ value the reliability of getting their staple gluten-free items, like crackers, bread, and flour, without worry:

> “If there’s no hiccups, you know, I get my crackers, my bread, and sometimes flour. And I know I can get that.” (P09_Prescription User)

Finally, **convenience and support from prescription services** enhance the overall experience. Some participants benefit from the ease of ordering gluten-free products directly from their local pharmacy, which often includes additional services such as notifications when items are ready for pickup:

> “Well, because I don’t have to go to a supermarket…I can order it direct from my local pharmacy, they deal with it for me…I find that quite easy to order gluten-free food on prescription. And they text me when it’s there as well, so I don’t have to keep looking in to see if they’ve got it.” (P09_Prescription User)

##### 4.2.1.2 Perceived disadvantages

Participants also highlighted some disadvantages and challenges when using prescriptions to access GFF.

The **prescription ordering process** can be difficult for some individuals, with some describing it as cumbersome and time-consuming:

> “The fact you can’t get hold of it and you’ve gotta go out of your way to, to try and get there, taking hours out of your day chasing it around. It’s just, it doesn’t work, it’s not smooth, it’s not simple.” (P02_Caring for Child_Prescription User)

**Unreliability** in product availability was also a disadvantage, which included delays in receiving items due to supply issues. This resulted in incomplete orders and multiple trips, to the pharmacy if products were missing:

> “And sometimes they haven’t got all the products you’ve asked for…they give you a receipt and say go back when they’ve got it and you ring up a couple of days later, have you got this yet? “No”. And ring up a couple of days later, has this come in yet? “No”, or “yes”, and then you go down again. So months involves two trips instead of one trip.” (P13_Prescription User).

The **inconvenience of separate physical visits** to pharmacies also posed a challenge for some, especially for those with busy schedules or work commitments:

> “…for me, picking prescriptions up, it’s not as easy as walking into a supermarket. Supermarkets are open, you know, 8:00 until 10:00, and they’re open seven days a week. I think our pharmacy in Brecon is only open until midday on a Saturday. So, if you’re working, then that’s more difficult to pick up a prescription.” (P27_Sole purchaser)

The **quality of food** provided through prescriptions was also raised, with some participants dissatisfied with the taste and quality of the products:

> “I’m not going to lie, a lot of gluten-free food tastes crappy, and it seems like a lot of times we’re given the crappy ones with the prescription. And I’m like, okay, I’m thankful for the prescription, but this tastes horrible.” (P01_Card User).

At a societal level, participants were concerned about the **cost-burden** of prescriptions to the NHS:

> “I would say the cost to the NHS. It must be massive. It must be huge. The cost of dispensing is massive and I’m sure they’re paying a lot more for it than we would pay in the shops.” (P12_Prescription User).

**Food waste** was also a problem, with participants reporting issues when they received products they didn’t like or couldn’t use:

> “If you don’t like any of those brands or don’t get on with them then that’s it really. And not being able to try them, so you could try it and then not like it and then it’s then the hassle of trying to get that prescription changed and then it’s kind of then a waste really of… you could literally have one slice and go, this is horrible, and then literally end up having to throw, throw the things out really.” (P14_Sole purchaser).

##### 4.2.1.3 Suggested improvements

Participants suggested several changes to improve GFF access through prescriptions. One key suggestion was **improved child inclusivity**. Participants felt that current offerings via prescription are more suited to adults, leaving out options that might be more appealing or appropriate for children:

> “So, obviously, kids are not that interested in cream crackers. They would prefer—he’d prefer, you know, the jammy dodgers or something like that. So, I do feel it’s catered more towards adults—more fun, interesting things for kids and just for a more balanced diet and to be the same as his friend.” (P19_Caring for Child_Prescription User).

**Improving the ability to change orders** was also highlighted as a necessary improvement. Participants experienced frustrations with having to deal with unavailable products and inflexible prescription systems:

> “They changed the product, gave me a new prescription. And then I had a call. When I went to the pharmacy, they said, “oh, that product is not available. But we’ve got this product available”, which was the original one that I tried to get! So, I missed about three months where I didn’t have the product because I was back and forth the GP changing the product name on the prescription” (P24_Prescription User).

Improving **choice regarding the type and quantity of product** was another recommended improvement. Allowing individuals to choose their preferred products and quantities would enhance their overall experience:

> “Having that kind of independence to choose what I want and when I want it would make quite a big difference.” (P11_Prescription User).

**Process improvements** were also suggested, such as the ability to order repeat prescriptions online in pharmacies who do not yet offer this service to streamline the process.

#### 4.2.2 Subsidy card – views of pilot scheme service users

The four participants living within Hywel Dda Health Board who had signed up to the subsidy card pilot scheme reflected on their experiences of being part of the scheme.

##### 4.2.2.1 Experiences of moving from prescriptions to the subsidy-card

Most participants expressed **contentment with the transition** to the subsidy card.

> “As someone who has experienced the prescriptions in different places before I started using the pre-paid card, I can tell you the pre-paid card has been a lifesaver.” (P01_Card User)

> “I would say over the five years it’s been really helpful… It’s made life a lot more convenient in terms of access that you get with it.” (P06_Card User)

> “Oh, yes. I wouldn’t go back. Yeah, I do like having the card.” (P25_Card User)

The process of obtaining and using the subsidy card was described as **straightforward**. One participant found the application process smooth:

> “The application was quite **seamless**. It wasn’t as cumbersome as I thought it would be.” (P01_Card User)

Another highlighted the **ease of managing the card**:

> “Very straightforward. I can check my balance easily, and I receive updates when it’s been topped up.” (P17_Card User)

##### 4.2.2.2 Perceived advantages

The **flexibility and freedom** provided by the subsidy card were highlighted as the most beneficial aspects. One participant appreciated the increased control it offered them:

> “The most beneficial aspect, it’s the flexibility and the freedom, you know, that, that control aspect that I have.” (P06_Card User)

Others found the **variety of products available** to be a **significant improvement**:

> “The variety of things that I can get now, definitely, yes, it’s nicer, nicer alternatives than prescription ones.” (P25_Card User)

> “I’m just really thankful for the card. It gives me more leeway and more options.” (P01_Card User)

> “Generally, yeah, it is relatively easy… they do have a reasonable range of gluten-free food in there.” (P17_Card User)

Some participants particularly valued being able to use the card for more **specialised products** in store and online:

> “With the card you have options of what you want to pick… I can go to, like, artisanal bakeries, who’ll have stuff for there for people who have coeliac disease. And they’re really nice tasting.” (P01_Card User)

> “It gives me the scope to buy things that perhaps I wouldn’t… like teff flour and those sorts of non-wheat based flours which I can’t more easily get here.” (P17_Card User)

One participant described increased carbohydrate intake due **to the increased range of GFF available**, also allowing for more experimentation in cooking and variation in diet:

> “I would say there has been an increase in my carbohydrate intake…I also have enough access to gluten-free produce that I can use to make my own food and explore and experiment with recipes.” (P01_Card User)

They also found that the subsidy card makes **travelling away from home easier** by allowing them to purchase GFFs en-route, and felt that it **enhanced social interactions and feelings of inclusion:**

> “It makes travelling a little bit easier because you’re able to get gluten-free food without having to pack your bread and cereal.” (P01_Card User)

> “It’s a lot easier now. And I, I know this may seem unconventional, but the card, you know, really has improved my social interaction.” (P01_Card User)

The **financial burden** of GFF is a significant concern for many, with coeliac disease but one participant felt that the subsidy card has helped to alleviate this:

> “I would say it’s definitely a lot more affordable [using the subsidy card]. It, it gives me a lot less of a headache these days.” (P01_Card User)

However, another participant noted that while the card offers additional purchasing options, it has not drastically changed their overall financial situation:

> “I’m lucky that… I wouldn’t be going without things if I didn’t have the card.” (P17_Card User)

##### 4.2.2.3 Perceived challenges

Whilst largely positive about their experiences, some participants identified several challenges with the subsidy card, which could be improved.

Some participants have experienced challenges when **checking the card balance**:

> “To find out how much balance I’ve got left on the card, I do have to ring up, and it’s quite a process to go through with numbers.” (P25_Card User)

**Online card restrictions** were a challenge for some participants, when those online stores can offer a wide variety of products available.

> “It’s tricky with the online stuff… I know we can’t use it for Amazon, even though Amazon provide lots of nice healthy gluten-free food.” (P06_Card User)

Another participant experienced **issues with specific low-cost retailers**:

> “Home Bargains… offer really good prices on the things I buy, but my card isn’t accepted in there because it’s classed as a general merchandiser.” (P25_Card User)

Some participants faced difficulties regarding **the reliability of the availability of products in supermarkets**:

> “The supermarkets have a habit of stocking the new things for a while and then randomly just taking them away because they’ve decided they’re not selling enough.” (P06_Card User)

Whilst most participants felt that that the subsidy card offered ease of access to a greater range of GFF, they reflected that there are **geographical limitations** to this, with those in **rural areas** less likely to see the same benefits:

> “The card would be less effective for places that don’t have gluten-free options… Rural areas won’t enjoy the benefit of the card as much as someone who’s in an urban area.” (P01_Card User)

> “I think the card is quite flexible… But where I’m living geographically, I’m very limited to the options that are available in the shops.” (P06_Card User)

> “Not particularly locally because I live in [small Welsh town], which is quite a small village… my best chance is to go to a larger supermarket… for a wider choice of food.” (P25_Card User)

> “If I need to travel to [larger Welsh town], which is about 18 miles from here, to get a really good range.” (P25_Card User)

Another participant also highlighted the **challenges** of finding **culturally specific gluten-free options**, particularly for traditional dishes:

> “Having coeliac disease is harder when you’re part of an ethnic minority… sometimes you just want something that, you know, tastes like home.” (P01_Card User)

Whilst these processes have now changed, it is worth noting that one participant experienced challenges with separating out their groceries and the reporting requirements during the **early phase of the pilot**:

> “When you go to the supermarket, I always have to separate the stuff that has to do with gluten, so I use the card for that separately.” (P01_Card User)

> “I had to report it was stolen and I’m also being asked to submit receipts… to ensure that you’re consistently using it for gluten products.” (P01_Card User)

#### 4.2.3 Subsidy card - views of service users in other health boards

We explored the views of participants in other health boards on the subsidy card scheme. Specifically, they expressed the perceived advantages, potential concerns, and their views on switching should the scheme become available.

##### 4.2.3.1 Perceived advantages of subsidy card

Participants not yet on the subsidy card scheme identified several perceived advantages, highlighting its potential to improve various aspects of their experience.

**Increased choice** was perceived as a significant benefit for many. Participants valued the potential to choose from a wide range of gluten-free products without the restrictions they currently experience with the prescriptions:

> “So my hope would be that I would be able to choose the products that I want. And that there wouldn’t be a limit on you can only use it for bread or cereal or biscuits or whatever, that actually, there’s a range…you can use this against any gluten-free products in any shop.” (P26_Sole purchaser)

The subsidy card is also perceived to give patients’ **greater autonomy in managing their condition.**

> “I think a prepayment card that didn’t depend on it being prescribed in a pharmacy getting it in, so you had more control over what you got when. Because it also kind of it takes away freedom a bit if you’re being dictated to as to what you can have when and what you can eat and when you can eat by what someone can get in and when they can’t get it in.” (P23_Sole purchasers)

One participant who already complements their prescription items with items bought at the supermarket noted that it would afford them greater choice. Instead of choosing the cheaper option at the supermarket, which is what they do when they are self-paying, they could **choose what they prefer to eat:**

> “I think a pre-paid card would mean more variety again, because a lot of times I’ll go in and say if I’m buying like a loaf, I will choose what is cheaper, rather than which one I prefer. I think it’d give me a lot more options.” (P05_Prescription User)

Others felt that the increased choice available to them would **improve their diet**.

> “I’ll be having more of the product that I enjoy and ultimately eating better than what I am at the moment.” (P03_Prescription User)

The **convenience and time-saving benefits** of a subsidy card were also emphasised. Participants noted that being able to purchase gluten-free products directly from local stores, rather than dealing with the complexities of prescriptions, would be significantly more efficient:

> “I can pick up the Warburton’s gluten-free bread in my local Tesco’s, I can walk five minutes down the road and pick it up. But to try and get it on prescription has taken me two months and numerous trips into town. So, it just, it doesn’t work.” (P02_Caring for Child_Prescription User)

Some felt that the subsidy card would also provide **more holistic support system** that recognised their individual needs.

> “Yeah. I mean I have to say that if we went down this route [subsidy card], I would feel much more supported in terms of the holistic needs of a person with coeliac disease.” (P16_Caring for Child_Prescription User).

##### 4.2.3.2 Concerns of the subsidy card

In addition to the perceived advantages, some potential concerns were noted by participants.

One concern was the **value of the card within the broader economic climate.** Participants worried that the card might be impacted by inflation and price fluctuations, which could affect its value over time and location:

> “I suppose the disadvantage would be that it would be subjective to inflation and to the price changes and therefore may go further for some individuals than it does in others.” (P16_Caring for Child_Prescription User)

Another perceived issue was the **increased taxpayer burden.** Participants expressed concern that a subsidy card system might lead to higher costs for taxpayers, as more people with coeliac disease might use the card compared to the current prescription system:

> “If I’m being honest, you know, hand on heart, the disadvantage comes to the taxpayer. Because more people who are coeliac are likely to use a card, whereas fewer of us use the prescription offer. So I think the disbenefit is to the taxpayer.” (P26_Sole purchaser)

**Potential misuse** of the card was another concern. Participants worried about the possibility of improper use (e.g. purchasing non GFF items) or fraud, although they acknowledged that there could be mechanisms to mitigate these issues.

> “The disadvantage, I suppose, is if it wasn’t being used properly. But I mean there’s, there’s ways of controlling that, isn’t there? It can only be used to purchase certain items…” (P02_Caring for Child_Prescription User)

**Potential top-up failures** were also noted as a possible disadvantage. Participants feared that technical issues or failures in adding funds to the card could leave them without the necessary products:

> “Well only if the top up failed or something, if you were left there without anything on it when there should be stuff on it really. I mean that would be a disadvantage.” (P13_Prescription User)

Additional barriers were noted for those who live in **rural locations.** Participants valued the convenience of prescriptions being available without needing to travel far, which might not be as feasible with a subsidy card.

> “On the flip side I think it is good with the prescriptions that it’s just there and I have to go and get it. It can be a bit of a faff trying to access it [the process], but at the end of the day there is food for me that I don’t have to worry too much about having to travel too far for. So, living rurally [the prescription] is quite good.” (P11_Prescription User)

Finally, there were some concerns **about shops’ familiarity with the card.** Participants expressed concerns that some stores might not accept the subsidy card or might not be familiar with the system, which could limit where they could use it.

> “Disadvantages would be if certain places didn’t accept it and that you were still restricted to certain locations to buy, you know, those products, which I think would still be fine.” (P07_Prescription User)

##### 4.2.3.3 Views on switching to a subsidy card

Participants had a range of views on switching to the subsidy card system. Sixteen (70%) participants expressed clear interest in switching to the subsidy card system, six (26%) were willing but ambivalent, and only one explicitly mentioned a lack of interest in switching.

Some participants **wanted to switch to the new scheme**, should it become available, noting that they would prefer it over the prescription model:

> “I mean obviously the card system…that sounds ideal. When I read about it, I was like, this seems like a really good idea and I’d much prefer that. So if I were to go into the shop and choose what I wanted and then you’d pay for it with a sort of discount on the product.” (P07_Prescription User)

> “The card I think is a fantastic idea…the dietician actually mentioned that and she said, ‘There’s other Health Boards doing that.’ I was like, ‘Oh that would work so much better.’ The current system just doesn’t work for anybody, it really doesn’t.” (P02_Caring for Child_Prescription User)

Others felt like it was a good idea, but they **required more information** about how the card is used before fully committing:

> “It sounds a good idea but how—how much is on that card? How…? What would be the discount? And Is it loaded with so much money and then it gets knocked off each time you use it, or…? Probably, yes, if I got more information, you know, written down and just read it through. But it sounds a good idea.” (P08_Prescription User)

> “Well, I’d look at, you know, what terms, conditions and things. But yes, possibly.” (P21_Prescription User)

Some wanted **specific reassurance** that they would not be at a **financial disadvantage**:

> “Depends how much it’s loaded with.” (P22_Prescription User)

> “Well, I would want to compare the value of what they’re offering me per month on a card as to what I can get on prescription.”(P09_Prescription User)

**Opposition to the subsidy card** was expressed by one participant who cares for someone who is a prescription user. They preferred the current prescription system or had reservations about potential changes:

> “Let me be honest, I won’t really accept it. I won’t.” (P10_Caring for Spouse_Prescription User)

> “Well really, to me, that’s – as opposed to what I can get on prescription for nothing-14 pounds isn’t going to get far… especially, say if I was disabled and I can’t get to a big supermarket and I’d only got my little supermarket here, 14 pounds for a month when it’s three pounds plus for a-just for a loaf-“ (P09_Prescription User)

#### 4.2.4 Experiences of coeliac disease health care services in Wales

Participants described their coeliac disease health care experiences. Some participants were satisfied with their care and felt that they had the **necessary access to support** when required.

> “I used to have a once a yearly review with a dietitian and now it’s just if I feel I need a chat with them, I can ring them up and ask… I feel I’m fairly well covered with it.” (P25_Card User)

> “Yeah, we get an annual review, yeah, that’s not a problem, from the dieticians” (P12_Prescription User)

One participant reported **very positive experiences**:

> “Absolutely amazing. I think, you know, I just think they are second to none. And they’ve been very helpful. They’ve been very clear with the information and support that they’ve provided. I just think that they’re brilliant” (P26_Sole purchaser)

Some participants felt that they had **limited interactions with health care professionals**. Some were happy with the set-up and **did not expect further interactions**:

> “But I don’t think we’ve ever had a coeliac disease review from the GP, nor would I expect one necessarily. You know she has teams in the hospital for that, so we don’t have a huge amount of contact with healthcare professionals over and above the receptionist team and the prescription clerk team, and they are all very understanding.” (P16_Caring for Child_Prescription User)

However, **others wanted more contact**. For example, one participant felt that coeliac disease is not a priority when compared to other illnesses:

> “Well, since diagnosis, I haven’t had any sort of follow-up appointment at all to see how I’m getting on” (P03_Prescription User)

> “And they didn’t really seem to prioritise it. I think they were just like, oh it’s food, whatever, you know, it’s not like actual medication. So I’ve really, really been unimpressed by that” (P07_Prescription User)

One participant acknowledged that the **pressures on GP services** likely contributed to the limited interactions, noting that they often had to remind their doctors about necessary check-ups:

> “I’m mindful that, you know, GP services are stretched… I have to remind them for my annual check-ups and things. I haven’t had a DEXA scan since I was first diagnosed.” (P17_Card User)

#### 4.2.5 General experiences

In addition to reflecting on the prescription and subsidy-card routes to obtaining GFF, participants also reflected more broadly on their general experiences of having coeliac disease, and the impact that has on their day to day lives.

##### 4.2.5.1 Variety of and access to GFF

Participants expressed varying levels of satisfaction with the range of gluten-free food available to them in supermarkets, with some acknowledging the more recent improvements. Most were **satisfied with the range** of gluten-free food available to them in supermarkets:

> “Yeah, so I think providing I shop in more than one place, I can get enough of the food that I want and that I need.” (P26_Sole purchaser)

> “But yeah, as far as buying foods go, I’ve got quite a good range around where I live. My only problem is it is very expensive.” (P28_Prescription User)

> “There’s quite a good range in the supermarkets. I find it seems to be getting a little bit better all the time.” (P24_Prescription User)

> “Most supermarkets now provide a good range.” (P08_Prescription User)

> “They’ve got a good selection, and they’ve always got the things in stock that I need. So, it’s a lot quicker when time is of the essence.” (P19_Caring for Child_Prescription User)

However, some found it **challenging to access the variety** they wanted, especially if they lived in more **rural** locations:

> “I live in quite a rural location. And so my choice of supermarkets is quite limited. I tend to shop for gluten-free things when I go further afield, but it is limited where I live. There’s no consistency, either, you know, one week, you might be able to buy the bread that you like, and then the next you can’t.” (P27_Sole purchaser)

> I would like to see a bigger range. But then, my sort of scope is restricted by the area I live in. If I lived in Cardiff, I’m sure I could find a greater variety.” (P27_Sole purchaser)

Others reported the inconvenience of **multiple store visits and additional travel required** to obtain the desired variety of GFF, adding significant burden to their daily routines:

> “So I can’t do a shop in one place. I know where to access what I need…but I’ve got to go out of my way. It is in my daily routine, or my weekly routine. But I’ve had to flex things to do it. It doesn’t slot in. I can’t do what other people do we just go in nip and buy something on the way home, it dictates my life.” (P23_Sole purchasers)

> “It adds on additional travel that I wouldn’t normally do…so it’s just very hard.” (P07_Prescription User)

Further, some were worried about the **cross-contamination risks** with supermarket products.

> “I’m always a little bit hesitant about the gluten free flour that you can buy on the supermarket, because in nine cases out of 10 it’s lumped in next door to something else that is liable to have cross-contamination.” (P15_Prescription User).

##### 4.2.5.2 Eating out

Participants also reflected on their experiences of eating out with coeliac disease. They report **challenges with eating out**, as finding gluten-free options at restaurants was often difficult.

> “As part of my job I drive around. Basically, whatever I want to eat that day, I have to take with me. So I can’t go and pick up a meal deal…That’s a nightmare. So in terms of being out and about no, I have to take everything with me.” (P23_Sole purchasers)

> “Eating out I am limited to probably two or three places in this local town that I know I can eat from.” (P06_Card User).

> “The gluten-free and vegetarian thing limits me… I can never have the cauliflower cheese because it’s been thickened with wheat flour.” (P17_Card User).

Even when such options were available, there were concerns about **cross-contamination** and whether the food was genuinely gluten-free:

> “And that’s a whole other area, isn’t it? Eating food out gluten-free? Because it’s the availability and the anxiety that perhaps possibly what you’re being provided with, is not up to the standard that you require it.” (P27_Sole purchaser)

Others called for **clearer labelling and more options.** Improved transparency in product information would help people to be certain they are selecting appropriate products:

> “I’d like there to be more of them and more clearly signed, because I have to look very carefully at the ingredients. Another reason why I’ve stuck with Waitrose is that they list the ingredient clearly. It’s not look at the back of the packet; it tells you exactly what the allergens are, whether there’s wheat in it, whether there’s malt, whether there’s barley, etcetera and so forth.” (P18_Prescription User)

##### 4.2.5.3 Financial impact of coeliac disease

Many participants found that the **cost of GFF was higher than regular food**, which placed a **strain on their budgets and impacted them financially**. For example, one participant noted:

> “We’ve noticed a massive difference in our, in our finances every month now. We’re having to buy my son gluten free food, it’s everything. The bread is like three times, four times the price of…even more than that actually because they’re half loaves. I mean even things like chicken nuggets, you know if he wants chicken nuggets as a treat you go and buy them and they’re like £4.50 for a pack of 12 rather than, you know, £3 for 42.” (P02_Caring for Child_Prescription User)

> “We’ve noticed a massive increase, not only that, the food cost of living and the cost of food going up… like normal spaghetti, 45p… Tesco’s gluten-free spaghetti 90p, like that’s double.” (P14_Sole purchaser)

Participants emphasised that the **high costs place a financial strain on families**, particularly those with **multiple coeliac members**, and stressed the importance of continued financial support through prescriptions:

> “I would hope that Hywel Dda and NHS don’t stop prescriptions—full stop—because we’ve already mentioned several times that it’s a financial burden. You know, not for me but for families with children that have coeliac disease.” (P09_Prescription User)

The expense also extended to **school meals**, where families had to bear the **additional cost of providing gluten-free lunches** despite eligibility for free school meals.

> “the paediatric specialist said in September to take her off school lunches completely [due to cross-contamination]. It’s not fair because she’s coeliac…entitled to free school meals at the moment. that has made it more difficult and it’s more expense because we have to provide lunch every day.” (P04_Caring for Child_Prescription User)

On the other hand, others noted that it was not affecting them personally, but still recognised that the costs were higher:

> “I mean I think it does cost more, but I know it’s not adversely impacting like my financial situation, like I’ve still got enough. It’s manageable. It’s fine, but I know, I know that they, they are more expensive. Like a loaf of bread might be £3 or £4, you know, whereas a regular loaf of bread will be like £1 or £2.” (P07_Prescription User)

##### 5.2.5.4 Lack of awareness and education of coeliac disease

More broadly, the participants felt that **better awareness and education** about coeliac disease was important.

> “Hopefully, I think the more people talk about it [the better]. I did a whole podcast for about an hour about a month ago, about being coeliac, which is on YouTube that went down very well.” (P29_Prescription User)

This in turn **could improve others understanding of the GFF needs** of people with coeliac disease, which **could help to improve GFF access when eating out**.

> “Absolutely, and we don’t eat out hardly ever because people don’t take it seriously.” (P04_Caring for Child_Prescription User)

Participants highlighted the **importance of educating others** on the condition’s seriousness, with some **experiencing dismissive attitudes toward their dietary restrictions**.

> “Suppose a wider education of-you know, that people understand. I mean, in church on a Sunday… I took my own gluten-free wafers and I heard somebody scoff. And you think, that’s not very nice. I’m sorry, do you want coeliac disease?” (P21_Prescription User)

This lack of understanding extends beyond dining out, **impacting daily routines**, as coeliac **patients must scrutinise every food item to avoid gluten**.

> “It’s just huge, every single thing you do, you can’t do without thinking about it. And it’s, yeah, it’s challenging. And it can be quite exhausting… it’s like a minefield. It has helped having the Coeliac UK, having the guidebooks, and I’ve got the app on my phone” (P28_Prescription User)

For some participants, the **lack of awareness even affects areas like school meals**, where **inadequate knowledge can lead to cross-contamination**, causing children to fall ill.

> “Although they’ve provided a gluten-free menu… she was being ill like every week. It’s definitely cross-contamination.” (P04_Caring for Child_Prescription User)

#### 4.2.6 Main priorities of people with coeliac disease

In addition to their views on prescriptions, the subsidy card, and general experiences of accessing GFF, participants also outlined six key priorities for improving access to GFF and better supporting people with coeliac disease.

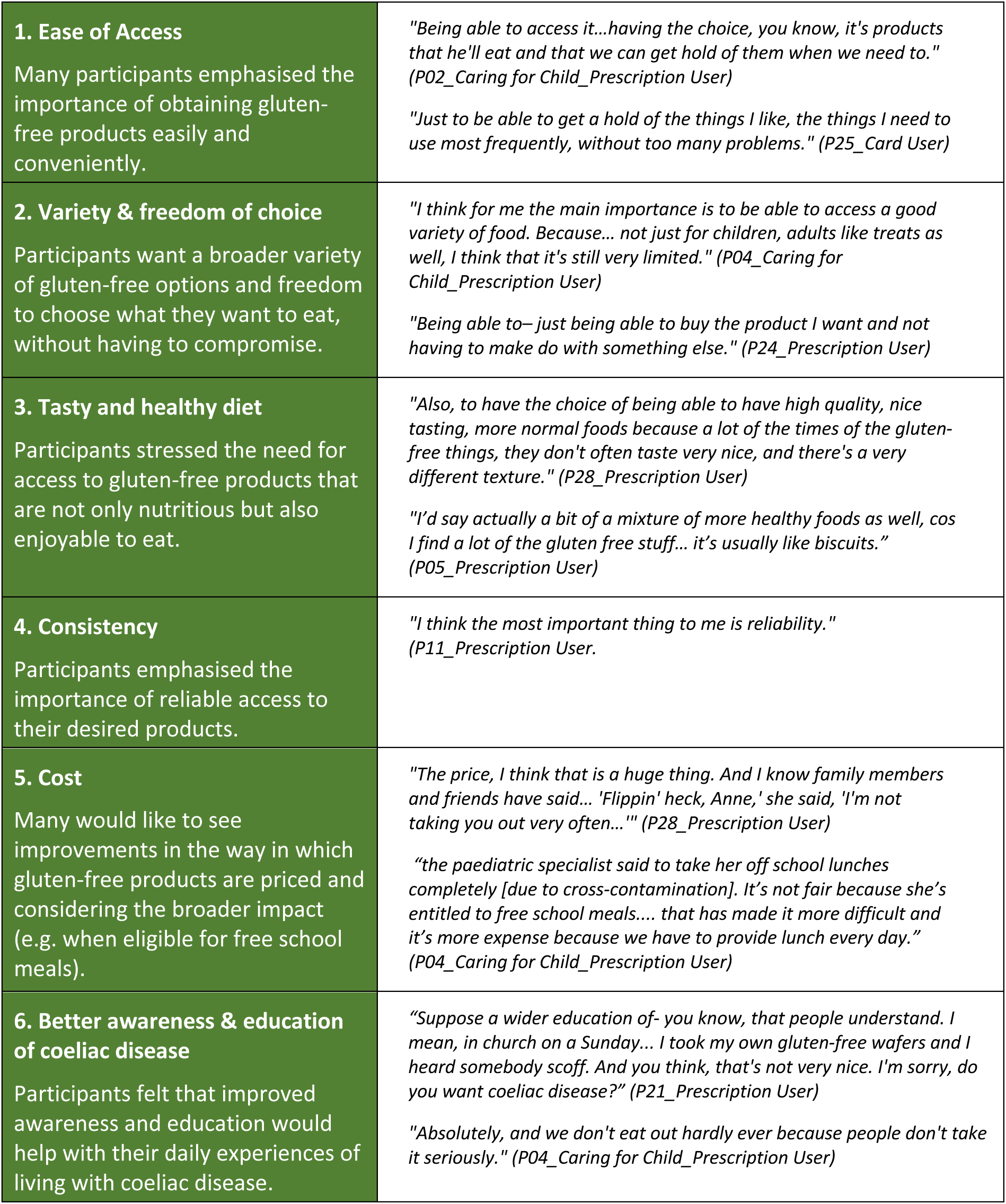

## 5. Discussion

We explored 29 service users’ views on the use of pre-paid subsidy cards in place of prescriptions, identified key barriers and facilitators to the subsidy card scheme, and highlight key recommendations for future roll-out. The majority of service users perceived multiple advantages of subsidy card scheme over the existing prescription system but did reflect on some potential concerns or challenges (see Box 2). Most people were positive towards the scheme −96% of participants (n=22) who are not part of the HDUHB pilot scheme expressed an interest in switching to the scheme should it become available.

Interviews also revealed participants’’ experiences of living with coeliac disease and six key priorities: 1) ease of access; 2) variety and freedom of choice; 3) tasty and healthy diet; 4) consistency; 5) cost; 6) better awareness and education of coeliac disease. Below we reflect on the key advantages, challenges, the potential of the scheme to address the six priorities identified by service users and make recommendations for future roll-out.

### Box 2

**Key advantages and concerns/challenges of the subsidy card scheme**

| Box 2 – Key advantages and concerns/challenges of the subsidy card scheme |  |
| --- | --- |
| Key advantages | Key concerns / challenges |
| Improved variety and choice<br>Healthier diet<br>Diets that meet personal preferences and needs<br>Increased flexibility and freedom<br>Times-saving benefits<br>Convenience<br>Improved social inclusion<br>Increased control<br>More holistic support for coeliac disease | Economic concerns (card value)<br>Card balance management<br>Technical issues e.g. top up failures<br>Retailer acceptance<br>Misuse / fraud<br>Cultural dietary needs<br>Geographical location<br>Financial burden on taxpayers |

### 5.1 Key advantages of the subsidy card

Both current and potential users perceived multiple benefits of the subsidy card scheme, over the prescription scheme (see Box 2).

One of the most frequently cited advantages is the **increased choice** the subsidy card provides. Current users highlight the freedom to select from a broader range of gluten-free products as being particularly valuable, reporting it as a marked improvement over the more limited options available through the traditional prescription system. This included access to more specialised products that are not currently available. Potential users also cite increased choice as one of the most attractive elements of the scheme, allowing them to **tailor their diets according to personal preferences and needs**. The card also facilitates experimentation with new recipes and dietary variations, supporting **healthier diets** and enhancing their overall quality of life. All of this was thought to increase **feelings of control** over managing coeliac disease.

The **freedom and flexibility** to shop at multiple locations was another significant benefit. Both groups of participants felt that it was more **convenient** to pick up the gluten-free products at the supermarkets, avoiding additional trips to the pharmacy. This made them feel like coeliac disease could **fit into their daily lives better.** Further, **time-saving benefits** are also considered highly valuable, as the card could eliminate the frequent bureaucratic and process complexities of obtaining prescriptions. In turn, improved flexibility, convenience and time-saving all help to make GFF **more accessible.**

More broadly, the card also appears to support **increased inclusivity**, with some users reporting that it has improved their ability to **engage in social activities** without the usual worries about gluten-free food options. Additionally, some participants feel that the card could provide a **more holistic support system**, allowing them to take greater control of their dietary needs and overall health.

### 5.2 Potential concerns, challenges and solutions

Despite multiple clear advantages reported by both groups, participants did also highlight potential concerns about the card or challenges experienced (see Box 2).

Potential users raised concerns regarding the **card’s monetary value** in relation to inflation and price increases, which could reduce its purchasing power over time. Other concerns related to **card management issues**, such as the ease of checking the card’s balance or potential technical issues such as top-up failures. Some find the existing system for checking the balance inefficient and cumbersome and would welcome improvements e.g. a phone app to check card balance and next payment date.

Potential users reported **uncertainty over where they could use the card** as a potential barrier to using it, especially in rural areas. There are also **limitations on where the card can be used**, particularly online, where retailers such as Amazon (known for offering a wide variety of gluten-free products) do not accept the card. Additionally, low-cost retailers such as ‘Home Bargains’ are not included in the scheme, limiting users’ ability to access affordable gluten-free products. Some users from ethnic minorities also expressed difficulty in finding **culturally specific gluten-free products**, which limits the card’s utility for certain groups; although it is not clear whether the alternative prescription system could meet this need. **Potential misuse** of the card was another issue raised, with participants fearing that individuals might use it to purchase non-gluten-free products or the card could be stolen; although they acknowledged that appropriate controls could mitigate this risk

One of the most discussed concern or challenge experienced with the subsidy card related to **geographical location.** Participants acknowledged multiple benefits (e.g. improved variety, convenience, flexibility), but they felt it was important to note that **many of these benefits were location dependent** and might not be realised for people living in rural areas.

In addition, there was also a broader concern about the **potential financial burden on taxpayers**, with some participants worried that more people might use the subsidy card than the current prescription system, leading to higher overall costs.

Most of the concerns raised by service users about the subsidy card scheme, either perceived or through experiences, are addressable through **clear communication, improved infrastructure, and expanded partnerships.** We present some possible solutions and next steps in attempting to address the concerns and challenges in Box 3.

#### Box 3

**Potential solutions to perceived concerns and challenges**

| Box 3 – Potential solutions to perceived concerns and challenges |  |
| --- | --- |
| Perceived concerns / challenges | Potential solutions / next steps |
| **Economic concerns:** Potential concerns over the card's value due to inflation and increased cost of living | **Card value reviews:** Regularly review the card's value to address inflation and market fluctuations.<br>**Communication campaign:** Communicate the review period / process to service users and highlight the broader benefits of scheme. |
| **Card balance management:** Difficulties in checking card balance due to cumbersome and inefficient system. | **Improved digital tools:** Work with service-users to co-produce a user-friendly mobile application for easy balance checks and updates, for those who are comfortable with using this technology. |
| **Technical issues:** Problems with topping up the card or system failures. | **Reliable technical support:** Ensure reliable card top-up processes and offer clear guidance about how to access customer support for technical issues. |
| **Retailer acceptance:** Uncertainty on which retailers accept the card. Not all retailers, especially online or budget stores, accept the card, limiting product availability and lower cost alternatives. | **Expanded retailer network:** Regularly review the list of participating retailers, and consider the inclusion of lower-cost stores, whilst balancing issues of potential misuse.<br>**Communication campaign:** Communicate the list of stores that accept the card, ideally localised so that it is relevant and informative to the service user. This could help service users to plan their shopping trips more effectively. |
| **Cultural dietary needs:** Difficulty finding culturally specific gluten-free products. | **Cultural product partnerships:** Partner with additional retailers to offer culturally specific gluten-free options. |
| **Potential misuse:** Risk of card misuse for non-gluten-free items or the card being stolen. | **Audit and review** - appropriate use of the card will be set out in the documentation used in signing up for the card. The use of the card may be audited by NHS Wales. Patients should report the loss or theft of the card to NHS Wales to ensure that the card is cancelled |
| **Geographic limitations:** Rural areas may have fewer options and less benefit from the card compared to urban areas. | **Prescription continuity:** Continue the use prescriptions in areas of need, which could include more rural areas or for those with limited mobility. |

### 5.3 Does the subsidy scheme address key priorities of service users?

During the interviews, we identified six key priorities of people with coeliac disease. Based on the service-users views on the subsidy card obtained during the interviews, we reviewed whether the subsidy scheme has the potential to address these six key priorities. Four of the key priorities are directly addressed by the scheme; two could be addressed indirectly, or directly with further changes to the scheme’s administration and additional communication strategies.

|  |  |
| --- | --- |
| **1. Ease of Access**<br>Many participants emphasised the importance of obtaining gluten-free products easily and conveniently. | One of the key reported benefits of the subsidy card is improved and easier access to products<br>The card would make GFF access more convenient and it reduced the time needed to access it |
| **2. Variety & freedom of choice**<br>Participants want a broader variety of gluten-free options and freedom to choose what they want to eat, without having to compromise. | The most commonly cited benefit was increased variety of options available to choose from<br>Participants reported that the card allowed them to make dietary choices that met the personal needs and preferences |
| **3. Tasty and healthy diet**<br>Participants stressed the need for access to gluten-free products that are not only nutritious but also enjoyable to eat. | Participants felt that the improved choice offered by the card would lead to improved diet<br>Participants felt it also allowed them to explore a wider range of more flavourful and specialised products that they do not currently have access to |
| **4. Consistency**<br>Participants emphasised the importance of reliable access to their desired products. | It is likely that the increased flexibility of using the card at multiple locations would help to support this priority |
| **5. Cost**<br>Many would like to see improvements in the way in which gluten-free products are priced and considering the broader impact (e.g. when eligible for free school meals). | The card cannot directly address this priority, but the scheme could consider widening the number of retailers so that it includes the lower-cost retailers<br>Regular reviews and appropriate adjustments would directly address this priority |
| **6. Better awareness & education of coeliac disease**<br>Participants felt that improved awareness and education would help with their daily experiences of living with coeliac disease. | The card cannot directly address this priority. However, the card being used in shops and restaurants might bring the disease more into the public's awareness<br>An additional communication strategy to partners (e.g. around issues of cross-contamination) could help to address this priority directly |

### 5.4 Potential unintended consequences of changing from prescriptions to the subsidy cards

Whilst the participants were positive about the scheme and report many advantages, it is important to acknowledge that the transition from a prescription-based system to a subsidy card system might result in some potential unintended consequences that could affect both service users and the healthcare system.

One significant unintended consequence is the potential increase in the **overall cost burden** on the healthcare system. The subsidy card is intended to offer greater flexibility and choice in obtaining GFF, and also helps to overcome some of the key challenges noted by people using the prescription system; including those who do not utilise their prescriptions currently. These benefits could lead to **increased uptake** as users are no longer constrained by these challenges. Increased uptake via the subsidy card scheme might result in higher costs for the healthcare system. On the other hand, there could be cost savings associated with reducing the expenses of prescription handling and pharmacist dispensing, but these savings need to be carefully evaluated against the potentially increased product uptake.

**Rurality** introduces further complexity into the discussion. For individuals living in rural areas, the prescription system may currently offer a more reliable solution, as local stores may not always stock the specific gluten-free products needed. The perceived advantages of the subsidy card might be diminished if patients in rural locations must travel long distances to reach larger supermarkets that accept the card. In areas with limited retail options or where local stores are unfamiliar with the subsidy card system, patients might face difficulties accessing a comprehensive selection of gluten-free products. Conversely, patients who live near pharmacies with regular GFF supplies may find the prescription system more convenient and reliable. Therefore, the advantages of the subsidy card could be unevenly distributed based on geographic location.

Moreover, the shift from a prescription system to a subsidy card could disrupt established support structures for patients accustomed to the current system. Long-term users of the prescription system may experience confusion or anxiety about transitioning to a new system. As such, some feel that the subsidy card should be an optional alternative rather than a replacement for the prescription system. They emphasise the need for continuity of care and support, suggesting that a phased or voluntary approach might alleviate some of the concerns associated with such a significant change.

A move to less healthy diets could also be an unintended consequence. Some coeliac might use the card to purchase less healthy gluten-free foods that are not available through prescriptions. This could lead to dietary choices that do not align with optimal health management for individuals with coeliac disease, and which do not align with broader public health goals.

Before conducting our research, we hypothesised that some patients might perceive that they will lose valuable pharmacy support, either for coeliac disease or other health issues. However, this did not feature as a theme nor was it raised by the sample included in this study.

### 5.5 Strengths and limitations

This research captures a diverse range of participant experiences, offering valuable insights into the varied challenges faced by both coeliac patients and parents of coeliacs, including those in rural areas. The study involved participants with different experiences, including those on the prescription system, those eligible but choosing not to use prescriptions, individuals on the subsidy card scheme, and those who declined the card. The qualitative methods allowed for an in-depth exploration of participants’ needs and the underlying reasons for their preferences, providing a comprehensive view of the challenges and potential solutions in accessing GFF.

However, while the participant group was relatively diverse, there may be a potential bias in this self-selecting sample, primarily due to the digital methods used for recruitment and data collection. The reliance on online recruitment may have excluded individuals who are less digitally literate or lack internet access, as most participants identified themselves as comfortable with digital platforms (see section 4.1.5). Future work should also explore the views of those who do not typically self-select for research participation and also use offline methods for data collection. Further, given the importance of geographical location, it would be important to target people living in rural communicates to explore their views.

### 5.7 Conclusions

The subsidy card scheme offers multiple perceived benefits to service users, including increased choice, flexibility, convenience and control in accessing their gluten-free products. It also directly addressed most of the key priorities stated by people living with coeliac disease. There are some important concerns or existing challenges that need further attention; most could be address through clear communication, improved infrastructure, and expanded partnerships. However, some concerns require additional consideration to ensure the scheme does not increase inequalities for certain groups of the population. This includes the economic concerns over the card’s value in the current high inflation and cost of living context and exploring the potential for geographical disparities in scheme benefits for those living in rural areas.

## Data Availability

All data produced in the present study are available upon reasonable request to the authors

## Abbreviations

ABUHB: Aneurin Bevan University Health Board
BCUHB: Betsi Cadwaladr University Health Board
CD: Coeliac disease
CTMUHB: Cwm Taf Morgannwg University Health Board
CVUHB: Cardiff and Vale University Health Board
DM: Dermatitis Herpetiformis
GF: Gluten free
GFF: Gluten free foods
GP: General Practitioner
HDUHB: Hywel Dda University Health Board
I: Interviewer
NHS: National Health Service
P: Participant
PRIME Centre Wales: Wales Centre for Primary and Emergency Care Research
PTHB: Powys Teaching Health Board
SBUHB: Swansea Bay University Health Board
SMREC: School of Medicine Research Ethics Committee

## 7 Acknowledgements

We would like to thank Andrew Evans, Emma Williams, Alison Jones and Beti-Jane Ingram for their time, expertise and contributions. We would like to thank the service users who committed their time to provide their lived experiences of coeliac disease and to provide their views on the services they receive. We are also grateful to Coeliac UK for their support with participant recruitment.

